# Obesity is associated with increased pediatric dengue virus infection and disease: A 9-year cohort study in Managua, Nicaragua

**DOI:** 10.1101/2024.04.02.24305219

**Authors:** Reinaldo Mercado-Hernandez, Rachel Myers, Fausto Bustos Carillo, José Victor Zambrana, Brenda López, Nery Sanchez, Aubree Gordon, Angel Balmaseda, Guillermina Kuan, Eva Harris

## Abstract

**Background:** Obesity is on the rise globally in adults and children, including in tropical areas where diseases such as dengue have a substantial burden, particularly in children. Obesity impacts the risk of severe dengue disease; however, the impact on dengue virus (DENV) infection and dengue cases remains an open question.

**Methods:** We used 9 years of data from 5,940 children in the Pediatric Dengue Cohort Study in Nicaragua to examine whether pediatric obesity is associated with increased susceptibility to DENV infection and symptomatic presentation. Analysis was performed using Generalized Estimating Equations adjusted for age, sex, and pre-infection DENV antibody titers.

**Results:** From 2011 to 2019, children contributed 26,273 person-years of observation, and we observed an increase in the prevalence of overweight (from 12% to 17%) and obesity (from 7% to 13%). There were 1,682 DENV infections and 476 dengue cases in the study population. Compared to participants with normal weight, participants with obesity had higher odds of DENV infection (Adjusted Odds Ratio [aOR] 1.21, 95% confidence interval [CI] 1.03-1.42) and higher odds of dengue disease given infection (aOR 1.59, 95% CI 1.15-2.19). Children with obesity infected with DENV showed increased odds of presenting fever (aOR 1.46, 95% CI 1.05-2.02), headache (aOR 1.51, 95% CI 1.07-2.14), and rash (aOR 2.26, 95% CI 1.49-3.44) when compared with children with normal weight.

**Conclusions:** Our results indicate that obesity is associated with increased susceptibility to DENV infection and dengue cases in children, independently of age, sex, and pre-infection DENV antibody titers.

**Key points:** We describe a doubling in the prevalence of obesity in a cohort of 5,940 Nicaraguan children followed over 9 years. Children with obesity were more likely to be infected with dengue virus and had higher risk of developing dengue disease.

## INTRODUCTION

The four dengue virus serotypes (DENV1-4) cause the most prevalent mosquito-borne viral disease in humans, with an estimated 100 million infections and 50 million dengue cases annually. Although most DENV infections are clinically inapparent, those with dengue disease present with high fever, rash, headache, and muscle and joint pain, and some cases can progress to life-threatening disease [1,2]. Alarmingly, the incidence of dengue is increasing worldwide [3]. The complexity of DENV immunity further aggravates this situation; while serotype-specific anti-DENV antibodies are protective against subsequent disease, specific pre-existing intermediate levels of serotype-cross-reactive antibodies can increase the risk of severe disease by a different DENV serotype [4–6]. Consequently, the development of a tetravalent vaccine that confers protection from dengue caused by each of the four DENV serotypes has been challenging.

Further, non-immunological factors, such as body composition and metabolic health, are associated with risk of infectious diseases. In addition to the multiple noncommunicable diseases associated with obesity, obesity is characterized by low-grade chronic inflammation that negatively affects the immune system [7–9]. Consequently, obesity is associated with increased viral transmission, worse clinical outcomes, higher risk of hospitalization, and suboptimal immune responses elicited by infection or vaccination [9–12]. Pediatric obesity, whose worldwide prevalence increased 7-fold from 1975 to 2016, has been shown to negatively influence infectious disease outcomes [13]. A meta-analysis of multiple dengue studies revealed that obesity is associated with severe pediatric dengue, though individual reports are mixed [14,15]. Furthermore, greater body surface area and body weight were found to positively correlate with Zika and chikungunya virus infection, respectively [16,17]. Yet, to date, no study has reported an association between obesity and pediatric DENV infection or non-severe dengue. Non-severe dengue, known as breakbone fever, is very debilitating, with a large negative impact on human health and adverse economic consequences [1,18]. Given that obesity and DENV are both on the rise in children, we used 9 years of data from 5,940 children in the Pediatric Dengue Cohort Study (PDCS) in Managua, Nicaragua, to investigate whether obesity is associated with DENV infection, dengue disease, and individual dengue clinical manifestations in children.

## METHODS

### Ethics statement

The human subjects protocol of the PDCS was reviewed and approved by the Institutional Review Boards of the University of California, Berkeley (2010-09-2245), the University of Michigan (HUM00091606), and the Nicaraguan Ministry of Health (CIRE-09/03/07-008). Parents or legal guardians of all subjects provided written informed consent, and participants aged 6 years of age and older provided assent.

### Study design

We used a longitudinal analytical method to evaluate the association between obesity and DENV infection, dengue disease, and dengue clinical manifestations using data from the PDCS collected from February 2011 until April 2019. The PDCS is a prospective cohort study ongoing since 2004 conducted in the catchment area of the Health Center Sócrates Flores Vivas (HCSFV) in District 2 of Managua, Nicaragua, following ∼3,800 active children aged 2-17 years and collecting annual demographic data and healthy blood samples to detect arboviral infections [19]. The DENV inhibition enzyme-linked immunosorbent assay (iELISA) is used to measure anti-DENV antibodies in participants’ annual blood samples [5,20]. Clinically inapparent DENV infections are identified by seroconversion or a ≥4-fold increase in the DENV iELISA titer in two consecutive annual samples processed side-by-side in children who did not experience a laboratory-confirmed dengue case in the intervening year [19]. Dengue cases (clinically apparent DENV infections) are captured via enhanced passive surveillance at the HCSFV; acute and convalescent blood samples are collected from suspected dengue cases according to the 1997 World Health Organization (WHO) case definition and from undifferentiated febrile illnesses [19]. Dengue cases are confirmed by traditional or real-time reverse transcription-polymerase chain reaction (RT-PCR) [19,21,22], viral isolation, or seroconversion of DENV IgM antibodies [19,23] or seroconversion or a ≥4-fold increase in DENV iELISA titers in paired acute- and convalescent-phase samples [5,20,24]. Severe dengue disease is defined following the 1997 (Dengue Hemorrhagic Fever/Dengue Shock Syndrome; DHF/DSS) and 2009 (Severe Dengue) WHO guidelines [19]. Since 2011, the PDCS began collecting height and weight data; however, due to funding constraints, these data were not collected for all participants in 2013 and 2015.

### Data collection

All exposure variables (namely, height and weight, age, sex, and iELISA titer) were recorded during enrollment and yearly during the annual sampling (February-April). Height and weight measurements were collected in duplicate by a trained study staff, and if a >5% difference was identified, a third measurement was taken, and the two closest measurements were averaged. The outcomes of interest (i.e., DENV infection, dengue disease, and clinical manifestations) occur approximately 4-9 months after the annual sampling (peak dengue season is in August-December). Cases and clinical manifestations are recorded year-round for participants presenting with suspected dengue at the HCSFV.

### Body Mass Index z-score categories

Body mass index z-score (BMIz) was calculated using the ‘Analysis of Growth Data’ R package following the WHO protocol and WHO BMI reference population [25–27]. Participants were categorized into underweight, normal weight, overweight, or obese categories [13,26] (see Supplementary Material).

### DENV iELISA

The DENV iELISA is a competition ELISA that measures the inhibition of binding of a standardized polyclonal anti-DENV antibody conjugate to DENV1-4 antigens by each participant’s serum, as described previously [5,20]. DENV iELISA titers were binned into six categories; <10, 10-20, 21-80, 81-320, 321-1280, >1:1280 [4,5]. Here, iELISA titer <10 (i.e., DENV-naïve) represents the reference group.

### Statistical analysis

Logistic regression with generalized estimating equations (GEE; for robust standard error inference) was used to assess the association between obesity and the outcomes of interest. Two models were performed for each analysis: 1) an unadjusted model including BMIz category as the exposure together with the outcome of interest and 2) a multivariable model also including the participants’ sex and pre-infection age and DENV iELISA antibody titer as covariates. For the DENV infection analysis, a positive pre-infection DENV iELISA titer was used as a proxy for living in areas with higher risk for DENV infection. Further, given that participants with obesity are more likely to be infected with DENV (see results) – thus, more likely to have pre-existing DENV immunity, and considering that pre-infection DENV iELISA titers have been associated with dengue and severe dengue [4–6], we also adjusted for pre-infection DENV iELISA titers in our dengue (and clinical manifestation) analysis.

Additionally, different study populations were used depending on the analysis outcome. When studying associations with DENV infection, all participant observations were included in the analysis (i.e., all 5,940 participants; 26,273 person-years). However, two separate analyses were performed when studying associations with dengue disease: **A,** only including those infected with DENV (i.e., given DENV infection; 1,682 total infections) and **B,** including all participants regardless of DENV infection. All analyses for association with clinical manifestations only included participants infected with DENV. Statistical models, figures, and tables were generated using R studio version 2023.6.0.421 [28]. The ‘epiR’ package was used to calculate DENV infection incidence rate in all participants and the risk of dengue given DENV infection over the entire observation period [29]. The ‘geepack’ package was used to conduct the GEE, following an exchangeable correlation structure to account for repeated measurements of the same individual recorded annually [30]. The ‘gtsummary’ package was used to tabulate the results [31] and the ‘forester’ R package to create forest plot figures [32].

## RESULTS

During the study period of February 2011 until April 2019, a total of 33,676 observations (i.e., person-years) were recorded; however, only 26,274 (78.0%) data points from 5,940 unique participants with available height and weight data were included in our analysis. During the study period, we observed an increase in the prevalence of participants in the overweight (from 12% to 17%) and obese (from 7% to 13%) categories, a decrease in the normal weight category (from 79% to 68%), and no difference in the underweight category (Table 1). By 2019, 30% of the cohort was either overweight or obese. Overall, participants had a median age of eight years (IQR 5, 11); per BMIz category, eight (IQR 5, 11) for normal weight, ten (IQR 7, 13) for underweight, and ten (IQR 7, 12) for overweight and obese (Table 2). Among all participants, males and females were equally distributed; however, slight differences in sex distribution were observed by BMIz category, with males comprising 50% of normal weight, 57% of underweight, 48% of overweight, and 51% of obese observations (Table 2).

**Table 1.** Participants BMIz category by year.

| Year | BMIz Categories |  |  |  |  |
| --- | --- | --- | --- | --- | --- |
|  | Underweight | Normal weight | Overweight | Obese | Overall |
| 2011 | 59 (2%) | 2,357 (79%) | 365 (12%) | 209 (7%) | 2,990 |
| 2012 | 39 (1%) | 2,387 (78%) | 416 (14%) | 221 (7%) | 3,063 |
| 2013<sup>1</sup> | 18 (1%) | 931 (74%) | 187 (15%) | 116 (9%) | 1,252 |
| 2014 | 36 (1%) | 2,256 (74%) | 452 (15%) | 294 (10%) | 3,038 |
| 2015<sup>1</sup> | 45 (2%) | 1,581 (77%) | 246 (12%) | 180 (9%) | 2,052 |
| 2016 | 72 (2%) | 2,604 (74%) | 511 (14%) | 345 (10%) | 3,532 |
| 2017 | 62 (2%) | 2,626 (72%) | 567 (16%) | 378 (10%) | 3,633 |
| 2018 | 65 (2%) | 2,466 (72%) | 529 (15%) | 362 (11%) | 3,422 |
| 2019 | 81 (2%) | 2,247 (68%) | 546 (17%) | 417 (13%) | 3,291 |

<sup>1</sup> Height and weight data were not recorded for all participants in 2013 and 2015.

**Table 2.**
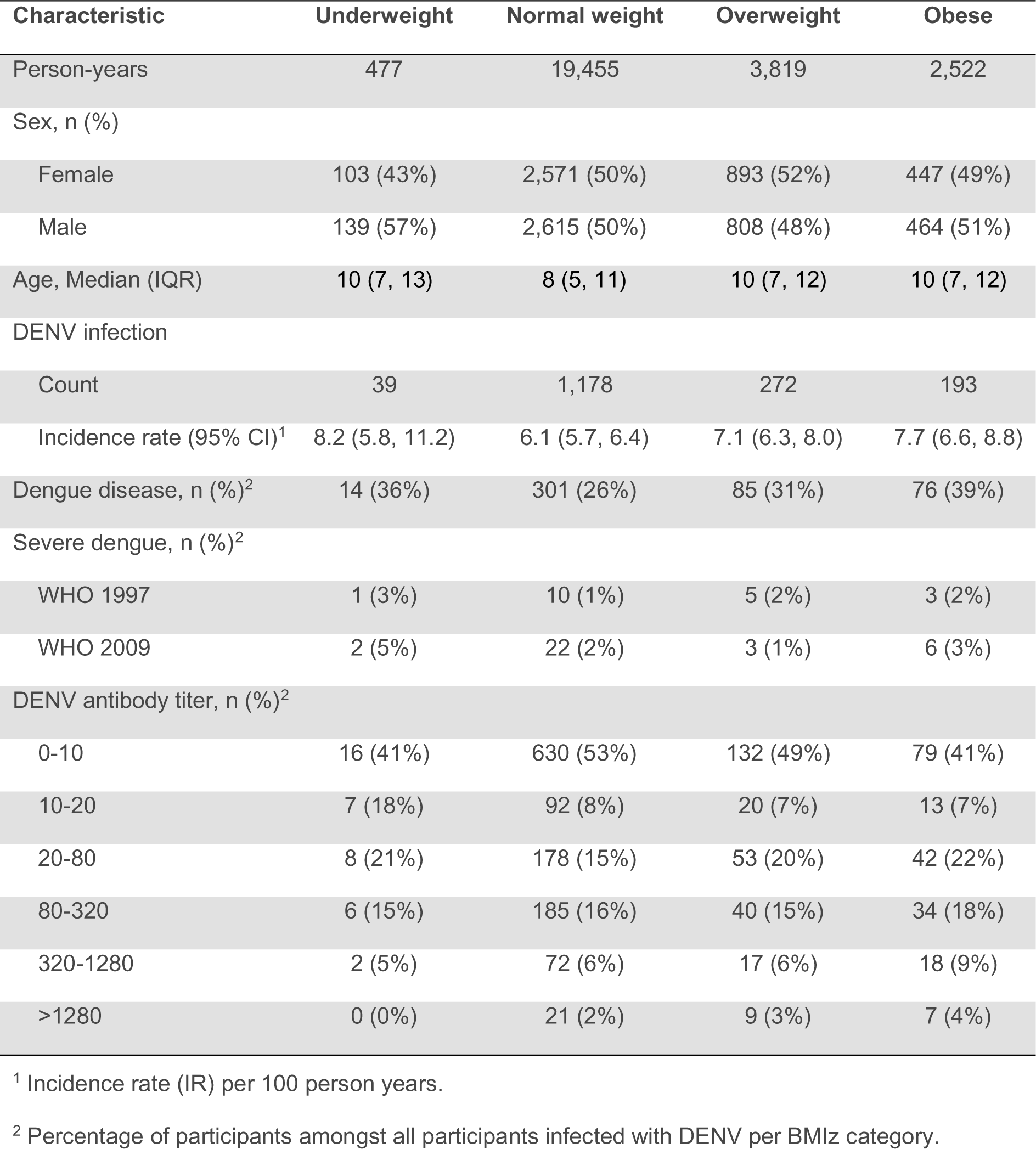
Participant demographics by BMIz category.

| Characteristic | Underweight | Normal weight | Overweight | Obese |
| --- | --- | --- | --- | --- |
| Person-years | 477 | 19,455 | 3,819 | 2,522 |
| Sex, n (%) |  |  |  |  |
| Female | 103 (43%) | 2,571 (50%) | 893 (52%) | 447 (49%) |
| Male | 139 (57%) | 2,615 (50%) | 808 (48%) | 464 (51%) |
| Age, Median (IQR) | 10 (7, 13) | 8 (5, 11) | 10 (7, 12) | 10 (7, 12) |
| DENV infection |  |  |  |  |
| Count | 39 | 1,178 | 272 | 193 |
| Incidence rate (95% CI)<sup>1</sup> | 8.2 (5.8, 11.2) | 6.1 (5.7, 6.4) | 7.1 (6.3, 8.0) | 7.7 (6.6, 8.8) |
| Dengue disease, n (%)<sup>2</sup> | 14 (36%) | 301 (26%) | 85 (31%) | 76 (39%) |
| Severe dengue, n (%)<sup>2</sup> |  |  |  |  |
| WHO 1997 | 1 (3%) | 10 (1%) | 5 (2%) | 3 (2%) |
| WHO 2009 | 2 (5%) | 22 (2%) | 3 (1%) | 6 (3%) |
| DENV antibody titer, n (%)<sup>2</sup> |  |  |  |  |
| 0-10 | 16 (41%) | 630 (53%) | 132 (49%) | 79 (41%) |
| 10-20 | 7 (18%) | 92 (8%) | 20 (7%) | 13 (7%) |
| 20-80 | 8 (21%) | 178 (15%) | 53 (20%) | 42 (22%) |
| 80-320 | 6 (15%) | 185 (16%) | 40 (15%) | 34 (18%) |
| 320-1280 | 2 (5%) | 72 (6%) | 17 (6%) | 18 (9%) |
| >1280 | 0 (0%) | 21 (2%) | 9 (3%) | 7 (4%) |

<sup>1</sup> Incidence rate (IR) per 100 person years.
<sup>2</sup> Percentage of participants amongst all participants infected with DENV per BMIz category.

During the study period, a total of 1,682 DENV infections were detected, resulting in an overall incidence rate of 6.40 (95% confidence interval [CI] 6.10, 6.72) per 100 person-years. In total, there were 476 dengue cases, with 19 meeting the 1997 WHO definition of DHF/DSS and 33 meeting the 2009 WHO Severe Dengue classification.

### Obesity is a risk factor for DENV infection in all PDCS participants

The BMIz-stratified incidence rate (per 100 person-years) of DENV infection was 6.05 (95% CI 5.71, 6.41) for normal weight, 8.18 (95% CI 5.81, 11.18) for underweight, 7.12 (95% CI 6.30, 8.02) for overweight, and 7.65 (95% CI 6.61, 8.81) for obese (Table 2). Using logistic regression GEE, we modeled the association between BMIz categories and odds of DENV infection over the entire observation period. In the unadjusted model, when compared with normal weight, participants in the overweight (odds ratio [OR] 1.19, 95% CI 1.04, 1.37) and obese (OR 1.30, 95% CI 1.11, 1.52) categories had significantly increased odds of DENV infection (Table S1). After adjusting for sex and pre-infection age and DENV iELISA antibody titers, only obesity was associated with increased odds of DENV infection (adjusted OR [aOR] 1.21, 95% CI 1.03-1.42; Figure 1A and Table S1). Furthermore, pre-infection DENV iELISA titers in the low and intermediate range were associated with higher odds of DENV infection (Figure 1A and Table S1). Conversely, titers higher than 1,280 were associated with lower odds of DENV infection (Figure 1A and Table S1).

**Figure 1.**
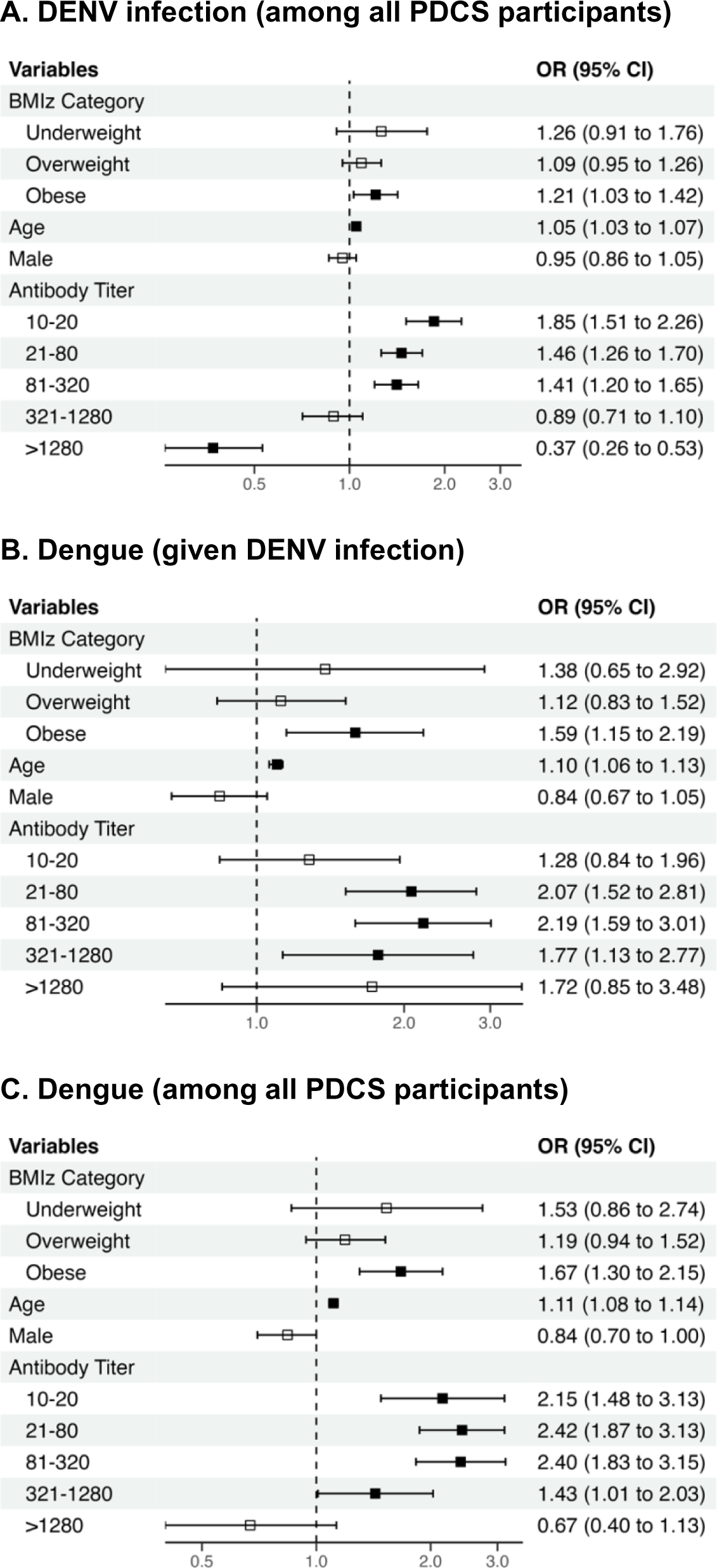
BMIz category and the odds of DENV infection, dengue disease given DENV infection, and dengue disease. *A*. BMIz category and the odds of DENV infection in all participants. *B*. BMIz category and the odds of dengue disease in participants infected with DENV (i.e., given DENV infection). *C*. BMIz category and the odds of dengue disease in all participants. Multivariable models are adjusted for sex and pre-infection age and DENV iELISA antibodies. Reference covariates: Normal weight for BMIz category, female for sex, and DENV iELISA titer <10 (i.e., DENV-naïve) for antibody titer. Odds ratio (square symbol) and 95% CI (horizontal line) are shown on each plot. Filled squares represent a statistically significant odds ratio. Abbreviations: OR, odds ratio; CI, confidence interval.

### Obesity is a risk factor for dengue disease in PDCS participants infected with DENV

To examine whether obesity was also associated with developing dengue disease in participants infected with DENV (i.e., dengue disease given DENV infection), we limited the analysis to the 1,444 children who experienced the 1,682 infections. Overall, the relationship between dengue disease among DENV infections followed a U-shaped curve, with the lowest proportion in children with normal weight (26%) and the highest proportion in children with obesity (39%; Table 2). In the logistic regression GEE model, only the obesity category was significantly associated with developing dengue in both the unadjusted (OR 1.89, 95% CI 1.39, 2.58) and the multivariable (aOR 1.59, 95% CI 1.15, 2.19) models adjusting for adjusting for sex and pre-infection age and DENV iELISA antibody titers (Figure 1B and Table S2). Furthermore, age (aOR 1.10, 95% CI 1.06, 1.13) was also associated with increased odds of developing dengue (Figure 1B and Table S2). Compared to DENV-naïve participants, DENV iELISA titers in the intermediate, but not the very low or very high, range were statistically associated with increased odds of dengue given infection (Figure 1B and Table S2).

### Obesity is associated with a greater risk of dengue in all PDCS participants

Next, we estimated the effect of obesity on dengue disease in all participants regardless of their DENV infection status. This analysis examines the association of BMIz category and the joint outcome of infection and symptomatic presentation among all PDCS participants, even uninfected participants who were not at risk of developing dengue. Given that capturing clinically inapparent DENV infections is not always possible without a prospective cohort study and serial serosurveys (as in the PDCS) this approach quantifies the association between BMIz category and dengue occurrence in a typical scenario where data on infection status is not available at the population level. In the unadjusted model, when compared with normal weight, participants with underweight (OR 1.88, 95% CI 1.06, 3.35), overweight (OR 1.44, 95% CI 1.14, 1.83), and obesity (OR 1.96, 95%CI 1.53, 2.52) had significantly increased odds of dengue (Table S3). Yet, in the multivariable model, only obesity was associated with higher odds of dengue (aOR 1.67, 95% CI 1.30, 2.15; Figure 1C and Table S3) when adjusting for sex and pre-infection age and DENV iELISA antibody titer. Age (aOR 1.11, 95% CI 1.08, 1.14) was also associated with increased odds of dengue and, compared to DENV-naïve participants, DENV iELISA titers in the low and intermediate range were associated with increased odds of dengue (Figure 1C and Table S3).

### Association of obesity with dengue clinical manifestations

To further examine the relationship between obesity and dengue disease, we next evaluated the odds of exhibiting individual dengue clinical manifestations among children with DENV infections. In multivariable models, compared to normal weight, obesity was associated with increased odds of fever (aOR 1.46, 95% CI 1.05, 2.02), headache (aOR 1.51, 95% CI 1.07, 2.14), and rash (aOR 2.26, 95% CI 1.49, 3.44) (Figure 2 and Tables S4-S6). No associations were observed between any of the BMIz categories and hemorrhagic manifestations, arthralgia, myalgia, leukopenia, or thrombocytopenia (Figure 2 and Tables S7-S11). Additionally, accounting for BMIz categories, pre-infection DENV iELISA titers in the intermediate range were associated with higher odds of presenting all the tested clinical manifestations, in most cases exhibiting a bell-shaped association curve (Table S4-S11).

**Figure 2.**
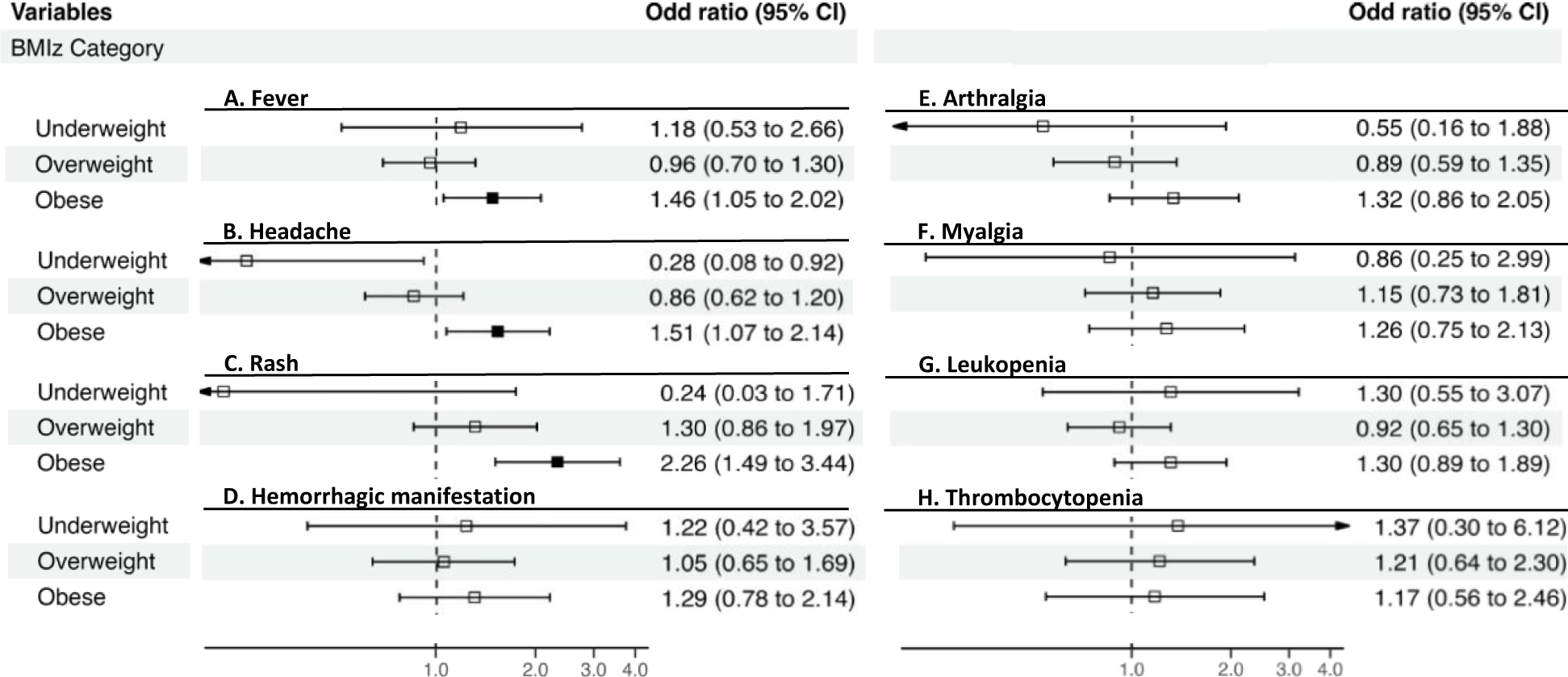
BMIz category and the odds of dengue clinical manifestations. BMIz category and the odds ratio of *A*, fever; *B*, headache; *C*, rash; *D*, hemorrhagic manifestation; *E*, arthralgia; *F*, myalgia; *G*, leukopenia; and *H*, thrombocytopenia in participants with DENV infection. Multivariable models are adjusted for age, sex, and pre-infection DENV antibodies. Reference covariates: Normal weight for BMIz category, female for sex, and DENV iELISA titer <10 (i.e., DENV-naïve) for antibody titer. Odds ratio (square symbol) and 95% CI (horizontal line) are shown on each plot. Filled squares represent a statistically significant odds ratio. Abbreviations: OR, odds ratio; CI, confidence interval.

## DISCUSSION

Here we report that obesity is associated with higher incidence of DENV infection and increased risk of dengue in a pediatric population, independently of pre-infection DENV antibody titer — a known risk factor for dengue disease [4–6,33]. The incidence of dengue is increasing worldwide; from 2000 to 2019, the number of dengue cases reported to the WHO increased by 10-fold [3]. Further, in 2019 and 2022, the PDCS in Managua, Nicaragua, experienced the largest dengue epidemics to date. Concurrently, the global prevalence of obesity has also been on the rise, including in the PDCS, which experienced a striking increase in the prevalence of overweight or obesity.

When observing all participants from 2011 until 2019, our analysis shows that obesity is an independent risk factor for DENV infection. These results are consistent with previous reports that mosquito biting frequency is positively correlated with individual weight or body surface area and that greater body weight or body surface area is associated with a higher prevalence of chikungunya virus or Zika virus infection, respectively [16,17,34].

In addition, when compared to normal weight, obesity was associated with increased odds of developing dengue disease in children infected with DENV. Specifically, we found that obesity was associated with higher odds of presenting fever, headache, and rash in children infected with DENV. Several studies have shown an association between obesity and a higher risk of severe dengue as compared to non-severe dengue [14,15,35–40]. However, we are not aware of reports of the association between obesity and non-severe dengue. Further, we confirmed previous studies from our group showing that pre-infection DENV iELISA antibody titers are associated in a bell-shaped curve with increased risk of developing dengue disease upon subsequent infection with DENV2 or DENV4, such that those with antibody titers in the intermediate range are at higher risk of dengue disease and severity [4,33]. By including both BMIz category and pre-infection antibody titer in our multivariable analysis, we demonstrate that obesity and pre-infection antibodies are both independent risk factors for developing dengue disease in children infected with DENV.

Given our prospective cohort study design, we were able to capture both clinically apparent and inapparent infections, thus permitting us to show that obesity is a risk factor for all DENV infection and that the association of obesity with dengue disease is independent from the risk of DENV infection. Since the odds of dengue in children with obesity will be driven by both risks, we conducted an additional, unrestricted analysis assessing the association between BMIz category and dengue including all PDCS participants regardless of DENV infection. The unrestricted model had slightly higher estimates (dengue in all PDCS participants, OR 1.67; vs dengue given DENV infection; OR 1.59). The larger estimate is the result of a mixed effect between the association with DENV infection plus the association with dengue disease. Further, the unrestricted analysis allows for comparison of our results with those from other studies or public health surveillance systems that rely only on clinically apparent infection and cannot control for differences in infection rate by BMIz categories.

Although we had good statistical power to test for the association between obesity and DENV infection (1,682 total infections) and subsequent development of dengue disease (476 dengue cases), one limitation was our low sample size to assess association with severe dengue (19 cases based on the 1997 and 33 based on the 2009 WHO classification). In this underpowered sample set, no significant association was observed between obesity and severe dengue among participants infected with DENV. Further, we were underpowered to detect associations with the underweight category (2% of participants were underweight; Table 1).

In conclusion, this study expands our current knowledge by demonstrating in a pediatric population that obesity is associated with DENV infection and is also associated with the development of dengue disease. Non-severe dengue is a very debilitating disease with enormous numbers of cases annually worldwide, major public health impact, and negative economic consequences, especially in low- and middle-income countries [18]. Given that people with obesity already experience a higher burden of noncommunicable diseases, an increased risk of DENV infection and disease add an extra layer of concern to public health. More work is needed to determine whether obesity is associated with increased DENV viral load and higher transmissibility of DENV from humans to mosquitoes, how obesity influences DENV evolution and viral fitness, and how it impacts the immune response post-DENV natural infection or vaccination. Our results, as well as others, suggest that infectious disease and immunological research, including studies of correlate of protection or disease risk, as well as vaccine trials, must consider non-immunological factors, such as body composition, in their analysis on infection rate, development of disease, immune responses, and vaccine efficacy.

## Supporting information

Mercado-Hernandez_2024_Supplemental files

## Data Availability

For data access arrangements, please contact E.H. at or the CPHS at. Standard data and material transfer agreements govern all materials and data used in this study.

## Authors contributions

R.M.H., F.B.C., and E.H. contributed to the conceptualization of the study. N.S., A.B., and G.K., supervised the cohort study. R.M.H., J.V.Z., B.L., and F.B.C. were responsible for data curation. R.M.H. and R.M. conducted the statistical analysis, with support from A.G. R.M.H. conducted the final analysis, figures, and tables. R.M.H. wrote the initial manuscript draft, and A.G. and E.H. edited the manuscript. R.M., F.B.C., J.V.Z., B.L., N.S., A.B., and G.K. reviewed the article for intellectual content.

## Acknowledgments

The authors thank our study personnel at the Sustainable Sciences Institute, the Laboratorio Nacional de Virología, and the Centro de Salud Sócrates Flores Vivas in Managua, Nicaragua, for their commitment and high-quality work and are grateful to all the children in the PDCS and their families.

## Data and materials availability

After securing approval from the UC Berkeley Committee for the Protection of Human Subjects (CPHS), individual data for figure reproduction can be shared with external researchers. For data access arrangements, please contact E.H. at or the CPHS at. Standard data and material transfer agreements govern all materials and data used in this study. The associated code is available for reference on GitHub.

