## Supplementary material for "Obesity is associated with increased pediatric dengue virus infection and disease: A 9-year cohort study in Managua, Nicaragua": Mercado-Hernandez_2024_Supplemental files

### Supplementary methods

#### BMIz category thresholds

For children younger than 5 years: **A)** underweight was defined as less than -2 BMIz, **B)** normal weight was defined as equal or greater than -2 BMIz and equal or less than 2, **C)** overweight was defined as BMIz greater than 2 but equal or less than 3, and **D)** obese was defined as a BMIz more than 3. For children 5-17 years old: **A)** underweight was defined as BMIz less than -2, **B)** normal weight was defined as BMIz equal or greater than -2 but equal or less than 1, **C)** overweight was defined as BMIz greater than 1 but equal or less than 2, and **D)** obese was defined as BMIz greater than 2.

### Supplementary tables

#### Table S1. Odds of DENV infection in all PDCS participants.

| Variable | PY | Event | OR[1] | 95% CI[2] | p-value |
|---|---|---|---|---|---|
| **Bivariable** | | | | | |
| **BMIz Category** | | | | | |
| Normal weight | 19454 | 1178 | 1.00 | (reference) | |
| Underweight | 477 | 39 | 1.38 | 0.99, 1.94 | 0.06 |
| Overweight | 3819 | 272 | 1.19 | 1.04, 1.37 | **0.012** |
| Obese | 2522 | 193 | 1.30 | 1.11, 1.52 | **0.001** |
| **Multivariable** | | | | | |
| **BMIz Category** | | | | | |
| Normal weight | 19356 | 1178 | 1.00 | (reference) | |
| Underweight | 476 | 39 | 1.26 | 0.91, 1.76 | 0.2 |
| Overweight | 3802 | 271 | 1.09 | 0.95, 1.26 | 0.2 |
| Obese | 2509 | 193 | 1.21 | 1.03, 1.42 | **0.019** |
| **Age** | 26143 | 1681 | 1.05 | 1.03, 1.07 | **<0.001** |
| **Sex** | | | | | |
| F | 13080 | 859 | 1.00 | (reference) | |
| M | 13063 | 822 | 0.95 | 0.86, 1.05 | 0.3 |
| **DENV iELISA antibody titer** | | | | | |
| 0-10 | 15635 | 857 | 1.00 | (reference) | |
| 10-20 | 1209 | 132 | 1.85 | 1.51, 2.26 | **<0.001** |
| 20-80 | 3154 | 281 | 1.46 | 1.26, 1.70 | **<0.001** |
| 80-320 | 3028 | 265 | 1.41 | 1.20, 1.65 | **<0.001** |
| 320-1280 | 1819 | 109 | 0.89 | 0.71, 1.10 | 0.3 |
| >1280 | 1298 | 37 | 0.37 | 0.26, 0.53 | **<0.001** |

[1] Odds Ratio (OR); [2]Confidence Interval (CI)

### Table S2. Odds of dengue in all PDCS participants infected with DENV

| Variable | PY | Event | OR[1] | 95% CI[2] | p-value |
|---|---|---|---|---|---|
| **Bivariable** | | | | | |
| **BMIz Category** | | | | | |
| Normal weight | 1178 | 301 | 1.00 | (reference) | |
| Underweight | 39 | 14 | 1.63 | 0.81, 3.29 | 0.2 |
| Overweight | 272 | 85 | 1.32 | 0.99, 1.77 | 0.058 |
| Obese | 193 | 76 | 1.89 | 1.39, 2.58 | **<0.001** |
| **Multivariable** | | | | | |
| **BMIz Category** | | | | | |
| Normal weight | 1178 | 301 | 1.00 | (reference) | — |
| Underweight | 39 | 14 | 1.38 | 0.65, 2.92 | 0.4 |
| Overweight | 271 | 85 | 1.12 | 0.83, 1.52 | 0.5 |
| Obese | 193 | 76 | 1.59 | 1.15, 2.19 | **0.005** |
| **Age** | 1681 | 476 | 1.10 | 1.06, 1.13 | **<0.001** |
| **Sex** | | | | | |
| F | 859 | 260 | Ref | (reference) | — |
| M | 822 | 216 | 0.84 | 0.67, 1.05 | 0.12 |
| **DENV iELISA antibody titer** | | | | | |
| 0-10 | 857 | 172 | Ref | (reference) | — |
| 10-20 | 132 | 36 | 1.28 | 0.84, 1.96 | **0.2** |
| 20-80 | 281 | 108 | 2.07 | 1.52, 2.81 | **<0.001** |
| 80-320 | 265 | 104 | 2.19 | 1.59, 3.01 | **<0.001** |
| 320-1280 | 109 | 41 | 1.77 | 1.13, 2.77 | 0.013 |
| >1280 | 37 | 15 | 1.72 | 0.85, 3.48 | **0.13** |

[1] Odds Ratio (OR); [2]Confidence Interval (CI)

### Table S3. Odds of dengue in all PDCS participants

| Variable | PY | Event | OR[1] | 95% CI[2] | p-value |
|---|---|---|---|---|---|
| **Bivariable** | | | | | |
| **BMIz Category** | | | | | |
| Normal weight | 19455 | 301 | 1.00 | (reference) | |
| Underweight | 477 | 14 | 1.88 | 1.06, 3.35 | 0.032 |
| Overweight | 3819 | 85 | 1.44 | 1.14, 1.83 | **0.003** |
| Obese | 2522 | 76 | 1.96 | 1.53, 2.52 | **<0.001** |
| **Multivariable** | | | | | |
| **BMIz Category** | | | | | |
| Normal weight | 19356 | 301 | 1.00 | (reference) | |
| Underweight | 476 | 14 | 1.53 | 0.86, 2.74 | 0.2 |
| Overweight | 3802 | 85 | 1.19 | 0.94, 1.52 | 0.2 |
| Obese | 2509 | 76 | 1.67 | 1.30, 2.15 | **<0.001** |
| **Age** | 26143 | 476 | 1.11 | 1.08, 1.14 | **<0.001** |
| **Sex** | | | | | |
| F | 13080 | 260 | 1.00 | (reference) | |
| M | 13063 | 216 | 0.84 | 0.70, 1.00 | 0.05 |
| **DENV iELISA antibody titer** | | | | | |
| 0-10 | 15635 | 172 | 1.00 | (reference) | |
| 10-20 | 1209 | 36 | 2.15 | 1.48, 3.13 | **<0.001** |
| 20-80 | 3154 | 108 | 2.42 | 1.87, 3.13 | **<0.001** |
| 80-320 | 3028 | 104 | 2.40 | 1.83, 3.15 | **<0.001** |
| 320-1280 | 1819 | 41 | 1.43 | 1.01, 2.03 | 0.047 |
| >1280 | 1298 | 15 | 0.67 | 0.40, 1.13 | **0.14** |

[1] Odds Ratio (OR); [2]Confidence Interval (CI)

### Table S4. Odds of fever in all PDCS participants infected with DENV

| Variable | PY | Event | OR[1] | 95% CI[2] | p-value |
|---|---|---|---|---|---|
| | | | **Bivariable** | | |
| **BMIz Category** | | | | | |
| Normal weight | 1178 | 288 | 1.00 | (reference) | |
| Underweight | 39 | 12 | 1.37 | 0.65, 2.92 | 0.4 |
| Overweight | 272 | 73 | 1.13 | 0.84, 1.53 | **0.4** |
| Obese | 193 | 70 | 1.76 | 1.29, 2.40 | **<0.001** |
| | | | **Multivariable** | | |
| **BMIz Category** | | | | | |
| Normal weight | 1178 | 288 | 1.00 | (reference) | |
| Underweight | 39 | 12 | 1.18 | 0.53, 2.66 | 0.7 |
| Overweight | 271 | 73 | 0.96 | 0.70, 1.30 | 0.8 |
| Obese | 193 | 70 | 1.46 | 1.05, 2.02 | **0.023** |
| **Age** | 1681 | 443 | 1.09 | 1.05, 1.13 | **<0.001** |
| **Sex** | | | | | |
| F | 859 | 236 | 1.00 | (reference) | |
| M | 822 | 207 | 0.90 | 0.72, 1.13 | 0.4 |
| **DENV iELISA antibody titer** | | | | | |
| 0-10 | 857 | 153 | 1.00 | (reference) | |
| 10-20 | 132 | 30 | 1.18 | 0.75, 1.86 | **0.5** |
| 20-80 | 281 | 105 | 2.33 | 1.71, 3.18 | **<0.001** |
| 80-320 | 265 | 99 | 2.38 | 1.72, 3.30 | **<0.001** |
| 320-1280 | 109 | 42 | 2.18 | 1.39, 3.41 | <0.001 |
| >1280 | 37 | 14 | 1.89 | 0.92, 3.85 | **0.082** |

[1] Odds Ratio (OR); [2]Confidence Interval (CI)

**Table S5. Odds of headache in all PDCS participants infected with DENV**

| Variable | PY | Event | OR[1] | 95% CI[2] | p-value |
|---|---|---|---|---|---|
| | | | **Bivariable** | | |
| **BMIz Category** | | | | | |
| Normal weight | 1178 | 228 | 1.00 | (reference) | |
| Underweight | 39 | 3 | 0.35 | 0.10, 1.16 | 0.086 |
| Overweight | 272 | 56 | 1.08 | 0.78, 1.49 | **0.6** |
| Obese | 193 | 60 | 1.88 | 1.35, 2.61 | **<0.001** |
| | | | **Multivariable** | | |
| **BMIz Category** | | | | | |
| Normal weight | 1178 | 228 | 1.00 | (reference) | |
| Underweight | 39 | 3 | 0.28 | 0.08, 0.92 | 0.036 |
| Overweight | 271 | 56 | 0.86 | 0.62, 1.20 | 0.4 |
| Obese | 193 | 60 | 1.51 | 1.07, 2.14 | **0.02** |
| **Age** | 1681 | 347 | 1.13 | 1.09, 1.18 | **<0.001** |
| **Sex** | | | | | |
| F | 859 | 192 | 1.00 | (reference) | |
| M | 822 | 155 | 0.80 | 0.63, 1.03 | 0.088 |
| **DENV iELISA antibody titer** | | | | | |
| 0-10 | 857 | 112 | 1.00 | (reference) | |
| 10-20 | 132 | 20 | 1.01 | 0.60, 1.68 | **>0.9** |
| 20-80 | 281 | 88 | 2.45 | 1.77, 3.41 | **<0.001** |
| 80-320 | 265 | 80 | 2.35 | 1.65, 3.36 | **<0.001** |
| 320-1280 | 109 | 36 | 2.22 | 1.37, 3.60 | 0.001 |
| >1280 | 37 | 11 | 1.57 | 0.74, 3.34 | **0.2** |

[1] Odds Ratio (OR); [2]Confidence Interval (CI)

### Table S6. Odds of rash in all PDCS participants infected with DENV

| Variable | PY | Event | OR[1] | 95% CI[2] | p-value |
|---|---|---|---|---|---|
| **Bivariable** | | | | | |
| **BMIz Category** | | | | | |
| Normal weight | 1178 | 109 | 1.00 | (reference) | |
| Underweight | 39 | 1 | 0.27 | 0.04, 1.91 | 0.2 |
| Overweight | 272 | 36 | 1.48 | 0.99, 2.23 | **0.058** |
| Obese | 193 | 40 | 2.54 | 1.70, 3.81 | **<0.001** |
| **Multivariable** | | | | | |
| **BMIz Category** | | | | | |
| Normal weight | 1178 | 109 | 1.00 | (reference) | |
| Underweight | 39 | 1 | 0.24 | 0.03, 1.71 | 0.2 |
| Overweight | 271 | 36 | 1.30 | 0.86, 1.97 | 0.2 |
| Obese | 193 | 40 | 2.26 | 1.49, 3.44 | **<0.001** |
| **Age** | 1681 | 186 | 1.07 | 1.02, 1.12 | **0.006** |
| **Sex** | | | | | |
| F | 859 | 106 | 1.00 | (reference) | |
| M | 822 | 80 | 0.77 | 0.56, 1.06 | 0.11 |
| **DENV iELISA antibody titer** | | | | | |
| 0-10 | 857 | 83 | 1.00 | (reference) | |
| 10-20 | 132 | 8 | 0.57 | 0.27, 1.18 | **0.13** |
| 20-80 | 281 | 47 | 1.60 | 1.07, 2.38 | **0.021** |
| 80-320 | 265 | 28 | 0.95 | 0.58, 1.54 | **0.8** |
| 320-1280 | 109 | 15 | 1.18 | 0.62, 2.24 | 0.6 |
| >1280 | 37 | 5 | 0.96 | 0.36, 2.50 | **>0.9** |

[1] Odds Ratio (OR); [2]Confidence Interval (CI)

**Table S7. Odds of hemorrhagic manifestations in all PDCS participants infected with DENV**

| Variable | PY | Event | OR[1] | 95% CI[2] | p-value |
|---|---|---|---|---|---|
| | | | **Bivariable** | | |
| **BMIz Category** | | | | | |
| Normal weight | 1178 | 89 | 1.00 | (reference) | |
| Underweight | 39 | 4 | 1.43 | 0.50, 4.11 | 0.5 |
| Overweight | 272 | 26 | 1.28 | 0.81, 2.04 | **0.3** |
| Obese | 193 | 22 | 1.57 | 0.96, 2.56 | **0.072** |
| | | | **Multivariable** | | |
| **BMIz Category** | | | | | |
| Normal weight | 1178 | 89 | 1.00 | (reference) | |
| Underweight | 39 | 4 | 1.22 | 0.42, 3.57 | 0.7 |
| Overweight | 271 | 26 | 1.05 | 0.65, 1.69 | 0.8 |
| Obese | 193 | 22 | 1.29 | 0.78, 2.14 | **0.3** |
| **Age** | 1681 | 141 | 1.11 | 1.05, 1.17 | **<0.001** |
| **Sex** | | | | | |
| F | 859 | 78 | 1.00 | (reference) | |
| M | 822 | 63 | 0.82 | 0.57, 1.18 | 0.3 |
| **DENV iELISA antibody titer** | | | | | |
| 0-10 | 857 | 50 | 1.00 | (reference) | |
| 10-20 | 132 | 10 | 1.16 | 0.57, 2.35 | **0.7** |
| 20-80 | 281 | 38 | 2.07 | 1.31, 3.26 | **0.002** |
| 80-320 | 265 | 22 | 1.21 | 0.70, 2.10 | **0.5** |
| 320-1280 | 109 | 15 | 1.89 | 0.98, 3.64 | 0.058 |
| >1280 | 37 | 6 | 1.89 | 0.71, 5.00 | **0.2** |

[1] Odds Ratio (OR); [2]Confidence Interval (CI)

### Table S8. Odds of arthralgia in all PDCS participants infected with DENV

| Variable | PY | Event | OR[1] | 95% CI[2] | p-value |
|---|---|---|---|---|---|
| **Bivariable** | | | | | |
| **BMIz Category** | | | | | |
| Normal weight | 1178 | 128 | 1.00 | (reference) | |
| Underweight | 39 | 3 | 0.67 | 0.20, 2.26 | 0.5 |
| Overweight | 272 | 31 | 1.07 | 0.71, 1.61 | **0.8** |
| Obese | 193 | 31 | 1.59 | 1.04, 2.44 | **0.033** |
| **Multivariable** | | | | | |
| **BMIz Category** | | | | | |
| Normal weight | 1178 | 128 | 1.00 | (reference) | |
| Underweight | 39 | 3 | 0.55 | 0.16, 1.88 | 0.3 |
| Overweight | 271 | 31 | 0.89 | 0.59, 1.35 | 0.6 |
| Obese | 193 | 31 | 1.32 | 0.86, 2.05 | **0.2** |
| **Age** | 1681 | 193 | 1.12 | 1.06, 1.17 | **<0.001** |
| **Sex** | | | | | |
| F | 859 | 109 | 1.00 | (reference) | |
| M | 822 | 84 | 0.80 | 0.59, 1.09 | 0.2 |
| **DENV iELISA antibody titer** | | | | | |
| 0-10 | 857 | 65 | 1.00 | (reference) | |
| 10-20 | 132 | 16 | 1.41 | 0.78, 2.52 | **0.3** |
| 20-80 | 281 | 41 | 1.65 | 1.07, 2.53 | **0.023** |
| 80-320 | 265 | 44 | 1.92 | 1.23, 2.99 | **0.004** |
| 320-1280 | 109 | 20 | 1.98 | 1.10, 3.57 | 0.023 |
| >1280 | 37 | 7 | 1.72 | 0.71, 4.14 | **0.2** |

[1] Odds Ratio (OR); [2]Confidence Interval (CI)

### Table S9. Odds of myalgia in all PDCS participants infected with DENV

| Variable | PY | Event | OR[1] | 95% CI[2] | p-value |
|---|---|---|---|---|---|
| **Bivariable** | | | | | |
| **BMIz Category** | | | | | |
| Normal weight | 1178 | 87 | 1.00 | (reference) | |
| Underweight | 39 | 3 | 1.06 | 0.32, 3.53 | >0.9 |
| Overweight | 272 | 29 | 1.50 | 0.96, 2.34 | **0.073** |
| Obese | 193 | 22 | 1.61 | 0.97, 2.65 | **0.064** |
| **Multivariable** | | | | | |
| **BMIz Category** | | | | | |
| Normal weight | 1178 | 87 | 1.00 | (reference) | |
| Underweight | 39 | 3 | 0.86 | 0.25, 2.99 | 0.8 |
| Overweight | 271 | 29 | 1.15 | 0.73, 1.81 | 0.5 |
| Obese | 193 | 22 | 1.26 | 0.75, 2.13 | **0.4** |
| **Age** | 1681 | 141 | 1.23 | 1.16, 1.31 | **<0.001** |
| **Sex** | | | | | |
| F | 859 | 81 | 1.00 | (reference) | |
| M | 822 | 60 | 0.79 | 0.55, 1.13 | 0.2 |
| **DENV iELISA antibody titer** | | | | | |
| 0-10 | 857 | 46 | 1.00 | (reference) | |
| 10-20 | 132 | 8 | 0.85 | 0.39, 1.83 | **0.7** |
| 20-80 | 281 | 36 | 1.76 | 1.11, 2.80 | **0.017** |
| 80-320 | 265 | 31 | 1.63 | 0.97, 2.73 | **0.067** |
| 320-1280 | 109 | 13 | 1.28 | 0.65, 2.54 | 0.5 |
| >1280 | 37 | 7 | 1.78 | 0.74, 4.26 | **0.2** |

[1] Odds Ratio (OR); [2]Confidence Interval (CI)

**Table S10. Odds of leukopenia in all PDCS participants infected with DENV**

| Variable | PY | Event | OR[1] | 95% CI[2] | p-value |
|---|---|---|---|---|---|
| | | | **Bivariable** | | |
| **BMIz Category** | | | | | |
| Normal weight | 1178 | 209 | 1.00 | (reference) | |
| Underweight | 39 | 10 | 1.60 | 0.72, 3.57 | 0.3 |
| Overweight | 272 | 54 | 1.15 | 0.82, 1.61 | **0.4** |
| Obese | 193 | 50 | 1.62 | 1.15, 2.30 | **0.006** |
| | | | **Multivariable** | | |
| **BMIz Category** | | | | | |
| Normal weight | 1178 | 209 | 1.00 | (reference) | |
| Underweight | 39 | 10 | 1.30 | 0.55, 3.07 | 0.6 |
| Overweight | 271 | 54 | 0.92 | 0.65, 1.30 | 0.6 |
| Obese | 193 | 50 | 1.30 | 0.89, 1.89 | **0.2** |
| **Age** | 1681 | 323 | 1.12 | 1.08, 1.17 | **<0.001** |
| **Sex** | | | | | |
| F | 859 | 187 | 1.00 | (reference) | |
| M | 822 | 136 | 0.72 | 0.56, 0.93 | 0.011 |
| **DENV iELISA antibody titer** | | | | | |
| 0-10 | 857 | 98 | 1.00 | (reference) | |
| 10-20 | 132 | 25 | 1.51 | 0.92, 2.49 | **0.1** |
| 20-80 | 281 | 83 | 2.66 | 1.89, 3.75 | **<0.001** |
| 80-320 | 265 | 80 | 2.75 | 1.92, 3.95 | **<0.001** |
| 320-1280 | 109 | 26 | 1.71 | 1.01, 2.90 | 0.047 |
| >1280 | 37 | 11 | 1.94 | 0.90, 4.18 | **0.092** |

[1] Odds Ratio (OR); [2]Confidence Interval (CI)

**Table S11. Odds of thrombocytopenia in all PDCS participants infected with DENV**

| Variable | PY | Event | OR[1] | 95% CI[2] | p-value |
|---|---|---|---|---|---|
| | | | **Bivariable** | | |
| **BMIz Category** | | | | | |
| Normal weight | 1178 | 39 | 1.00 | (reference) | |
| Underweight | 39 | 2 | 1.57 | 0.36, 6.83 | 0.5 |
| Overweight | 272 | 13 | 1.47 | 0.78, 2.78 | **0.2** |
| Obese | 193 | 10 | 1.59 | 0.78, 3.25 | **0.2** |
| | | | **Multivariable** | | |
| **BMIz Category** | | | | | |
| Normal weight | 1178 | 39 | 1.00 | (reference) | |
| Underweight | 39 | 2 | 1.37 | 0.30, 6.12 | 0.7 |
| Overweight | 271 | 13 | 1.21 | 0.64, 2.30 | 0.6 |
| Obese | 193 | 10 | 1.17 | 0.56, 2.46 | **0.7** |
| **Age** | 1681 | 64 | 1.07 | 0.98, 1.17 | **0.11** |
| **Sex** | | | | | |
| F | 859 | 35 | 1.00 | (reference) | |
| M | 822 | 29 | 0.85 | 0.51, 1.42 | 0.5 |
| **DENV iELISA antibody titer** | | | | | |
| 0-10 | 857 | 6 | 1.00 | (reference) | |
| 10-20 | 132 | 2 | 1.93 | 0.38, 9.65 | **0.4** |
| 20-80 | 281 | 25 | 11.90 | 4.74, 29.8 | **<0.001** |
| 80-320 | 265 | 19 | 9.51 | 3.62, 25.0 | **<0.001** |
| 320-1280 | 109 | 10 | 11.50 | 3.88, 34.2 | <0.001 |
| >1280 | 37 | 2 | 5.74 | 1.02, 32.3 | **0.048** |

[1] Odds Ratio (OR); [2]Confidence Interval (CI)
